# Concussion acutely disrupts auditory processing in Division I football student-athletes

**DOI:** 10.1101/2023.06.19.23291602

**Authors:** Jennifer Krizman, Danielle Colegrove, Jenna Cunningham, Silvia Bonacina, Trent Nicol, Matt Nerrie, Nina Kraus

## Abstract

Diagnosis, assessment, and management of sports-related concussion requires a multi-modal approach. Yet, currently, an objective assessment of auditory processing is not included. The auditory system is uniquely complex, relying on exquisite temporal precision to integrate signals across many synapses, connected by long axons. Given this complexity and precision, together with the fact that axons are highly susceptible to damage from mechanical force, we hypothesize that auditory processing is susceptible to concussive injury. We measured the frequency-following response (FFR), a scalp-recorded evoked potential that assesses processing of complex sound features, including pitch and phonetic identity. FFRs were obtained on male Division I Collegiate football players prior to contact practice to determine a pre-season baseline of auditory processing abilities, and again after sustaining a sports-related concussion. We predicted that concussion would decrease pitch and phonetic processing relative to the student-athlete’s preseason baseline. We found that pitch and phonetic encoding was smaller post-concussion. Student-athletes who sustained a second concussion showed similar declines after each injury. Auditory processing should be included in the multimodal assessment of sports-related concussion. Future studies that extend this work to other sports, other injuries (e.g., blast exposure), and to female athletes are needed.

## 1. Introduction

Public awareness of sports-related concussion is on the rise, and more athletes are being flagged for assessment following a suspected head injury; however, concussion is not always straightforward to assess, diagnose, and manage (1, 2). This difficulty is due in part because (a) a mild traumatic brain injury (mTBI), which is induced by an external mechanical force, affects function but not macro-structure (3, 4), (b) the symptoms linked to this injury are non-pathognomic (3, 5), and (c) in the case of athletes, oftentimes the ultimate goal of evaluating and treating the individual is to deem him or her ready to resume the activity that led to the injury (1, 6). Therefore, a multimodal approach has been increasingly recognized as a standard for concussion assessment, diagnosis, and management (1). Currently, this multimodal approach combines tests of cognitive function, sensorimotor abilities including oculomotor, vestibular, and coordination, and reports of somatic symptoms such as headache, dizziness, or nausea (1). While the recent inclusion of balance measures (i.e., BESS) and visual saccades (i.e., King Devick) offers some insights into the effects of head trauma on sensory (dys)function, inclusion of an objective auditory assessment could provide a more comprehensive picture of the pathophysiologic effects of head injury. In support of including an auditory assessment, concussed individuals often experience auditory complaints post-concussion, including ringing in the ears, an inability to ignore distracting sounds, and difficulty understanding speech in a noisy environment such as a restaurant or cafeteria (7-10).

There are structural and physiological reasons the auditory system merits inclusion in a neurosensory assessment following head injury. More relays connect the sensory organ to the brain than any other sensory system (11). Axons bi-directionally link these relays, traversing between the ear, brainstem, midbrain, and cortex (11-13). The precise signaling occurring along these multiple pathways requires a balance of neural inhibition and excitation choreographed across numerous neural synapses (14, 15) that work in both a feed-forward and feed-back direction (12, 13) to capture signals coming in at micro- to milli-second speeds(16). When a concussion occurs, axons are highly susceptible to damage from the mechanical force and are thought to be a chief site of dysfunction from traumatic brain injuries (3, 17-20). The long axonal tracts along the auditory neuraxis (11), comprising the lateral lemniscus lie perpendicular to the chief plane of concussion-causing force (21), making them a prime site for concussive damage. Shearing or stretching of axons as a consequence of a mechanical force can initiate a dysfunctional metabolic cascade (3, 5, 22) and, potentially, death of the injured axon (4, 17, 23-25). The susceptibility of axons in the auditory pathway to mechanical force together with the complex interconnectivity of the auditory system, makes the auditory system a prime site of dysfunction following a concussion.

There is accumulating evidence that sound processing is impacted by a brain injury. Humans with moderate to severe TBI (26, 27) and animal models with mild to moderate TBI show delayed or reduced neural responses to simple sounds such as a click or tone (28). Additionally, the frequency-following response (FFR), which measures responses originating predominately in the auditory midbrain (29, 30) and objectively assesses the processing of complex sounds, such as speech and music (16), has identified sound processing deficits in milder cases of TBI (8, 31-33). Specifically, the FFR shows delayed and reduced processing of speech sounds weeks after a sports-related concussion in adolescents with prolonged recovery (32) and months to years after recovering from a sports-related concussion in collegiate student-athletes (31). In adolescents with post-concussion syndrome, the effects observed include a reduction in encoding of the fundamental frequency (F0) (32). A decline in F0 amplitude has also been observed in collegiate student-athletes who have history of a single concussion and have ostensibly recovered from their injury (31) and in those who sustain a large number of subconcussive hits through participation in contact and collision sports (34). While these studies point to longer-term effects of sports-related concussion on sound processing, it is not known whether sound processing impairments are seen in the acute stage of concussion injury (i.e., 24-72 hours post-injury) or how this impairment manifests. Understanding the effects of sports-related concussion on auditory processing during the acute stage of injury is important because the FFR may provide a valuable adjunct in the clinical assessment of concussion. The FFR offers several advantages over other auditory assessments: it is highly reliable across test sessions – there are no practice effects, it is objective, meaning the patient does not need to respond in any way during the test, but may instead sleep or watch a movie, and the identical test can be used across ages and languages.

The long-term effects of concussion on speech processing reflect declines in processing the periodicity of lower-frequency, pitch-based cues. These pitch cues are important for identifying a speaker (e.g., knowing who is talking) and locking-on to that speaker’s voice in difficult or noisy listening conditions (35). However, pitch is not the only important speech feature that the brain needs to process. Speech also contains higher-frequency components that comprise formants, which determine the phonetic identity of the speech sounds (36). For example, this higher-frequency information distinguishes a ‘b’ from a ‘d’(36).

Given that the long-term deficits are specific to pitch encoding, one hypothesis is that pitch is selectively affected by a sports-related concussion. Alternatively, acute impairments following a concussion may be seen across all aspects of frequency encoding. That is, processing of both pitch *and phonetic* cues are disrupted acutely by a concussion but processing of phonetic cues recovers more quickly than processing of pitch cues. To arbitrate between these hypotheses, we collected FFRs on collegiate football players prior to the start of the football season to provide a baseline measure of auditory processing and tested concussed student-athletes again at 24-72 hours postinjury to identify acute changes in speech processing. If the pitch-specificity hypothesis is correct, then only impairments in encoding of the fundamental frequency (F0) the physical correlate of pitch perception, should be observed during the acute concussion phase. If the pervasive hypothesis is correct, then both pitchand phoneticencoding deficits should be seen acutely following a concussion.

## 2. Methods

### 2.1 Participants

Division I collegiate-football student-athletes (n=39 males, 19.51 ± 1.22 years of age at baseline testing) participated in this study. Student-athletes were administered FFR testing in the week prior to their first contact practice following study enrollment, providing a baseline to which responses collected after concussion were compared. The average number of days between baseline and concussion testing was 357.07 ± 354.94 and the average number of days between concussion diagnosis and testing was 2.81 ± 2.32. All concussions were football-related injuries. Thirty-three student-athletes sustained a single injury and six student-athletes sustained two concussions over the course of the study. Concussion diagnoses were made and recovery was managed by the team’s medical staff following the current NCAA concussion protocol(6). To participate in the study, participants must have normal peripheral auditory function, as indicated by normal distortion product otoacoustic emissions and a click-evoked auditory brainstem response peak V within normal limits. The sample size is sufficient to detect a concussion effect as small as η _p_ ^2^ = .14, indicating that we are well sampled to identify this effect. All participants consented to participate and study procedures were approved by the university’s Institutional Review Board in accordance with the Declaration of Helsinki.

### 2.2 Stimuli and Recording Parameters

Responses were elicited to a 40-ms ‘d’, presented in alternating polarity at a rate of 10.9 Hz. The ‘d’ is a five-formant synthesized speech sound (37) consisting of an initial noise burst and a formant transition between the consonant and vowel. The fundamental frequency (F0) and the first three formants (F1, F2, F3) change linearly (F0: 103-125, F1: 220-720, F2: 1700-1240, F3: 2580-2500 Hz) while F4 (3600 Hz) and F5 (4500 Hz) remain constant. The ‘d’ was presented through an insert earphone to the right ear at 80 dB SPL, which is a comfortable listening level. During data collection, the student-athlete sat in a comfortable chair and was instructed to rest or watch a captioned movie of his choice.

Frequency-following responses (FFRs) to the ‘d’ were collected using Ag/AgCl electrodes, with Cz referenced to the right ear lobe and forehead as ground. Impedance was kept ≤ 5 kΩ with interelectrode impedance < 3 kΩ. Stimuli were delivered and responses were collected with the Bio-logic Navigator Pro System (Natus Medical Incorporated, San Carlos, California). Responses were sampled at 12 kHz, filtered from 100-2000 Hz, epoched, and artifact rejected online. The epoch window was 75 ms long, beginning 15.8 ms prior to stimulus presentation. Two blocks of 3000 artifact-free trials were collected and responses were combined offline. The FFR responses were averaged in two ways to generate two 6000-sweep averaged responses: one that maximized the response to the fundamental frequency (F0), which predominantly reflects pitch encoding, and one that maximizes the response to the higher frequencies comprising the first and second formants (F1 and F2), which reflect phonetic encoding. To maximize the F0 response, the response to each polarity were added together and averaged, providing an FFR reflecting the stimulus envelope. The F1 and F2 responses were maximized by inverting responses to one of the polarities prior to averaging, thus emphasizing the response to the temporal fine structure (TFS)(38).

### 2.3 Data Analysis

#### 2.3a Measures of Spectral Encoding

Spectral encoding was analyzed using a fast Fourier analysis of the formant transition of ‘d’ (11 – 47.5 ms). Specifically, this segment was Hanning-ramped (2 ms on, 2 ms off), baselined to its mean amplitude to remove any DC component, and submitted to a 4096-point fast-Fourier transform (FFT). To determine F0 encoding, the spectral amplitude from 90-120 Hz was averaged. F1 was calculated by averaging the spectral amplitude from 200-740 Hz, and F2 was calculated by averaging the spectral amplitude from 1220 to 1720 Hz. Separate averaged amplitudes were extracted from the envelope and TFS responses.

#### 2.3b Statistical Analyses

Because the data were not normally distributed, spectral amplitudes were logtransformed prior to statistical analyses; however, all figures plot raw data for better visualization of the effects. Although the envelope maximizes the lower-frequency F0 and, to some extent, F1, and the TFS maximizes F1 and F2, all spectral amplitudes were analyzed together using a 2 (time: baseline, acute concussion) x 2 (polarity: envelope, TFS) x 3 frequency region (F0, F1, F2) RMANOVA, since energy from all frequency bins can still be meaningfully extracted from each polarity (38). We predicted, however, that the greatest effects for the F0 would be seen in the envelope response and the greatest effects for F1 and F2 would be seen in the TFS response. Planned comparisons were performed between the baseline and acute spectral amplitudes for each frequency region using paired-sample ttests. For the 6 student-athletes that sustained a second concussion, the same RMANOVA was run to compare their baseline FFRs to their second acute FFRs over F0, F1, and F2. Data processing was performed using custom routines coded in Matlab 2021a (The MathWorks, Inc., Natick, MA) and statistical analyses were performed in SPSS v29 (SPSS Inc., Chicago, IL).

## 3. Results

Acutely following a concussion, pervasive declines in frequency encoding are present (F_(1, 38)_ = 17.073, p < .001, η _p ^2^_ = 0.310, Table 1; Fig 1). As described below, this includes decreased processing of the F0, the speech component underlying pitch perception, and the formant frequencies (F1 and F2), which provide information about the phonetic identity of the speech sound (e.g., what distinguishes a ‘ba’ vs. ‘da’ or ‘da’ vs. ‘du’).

**Figure 1.**
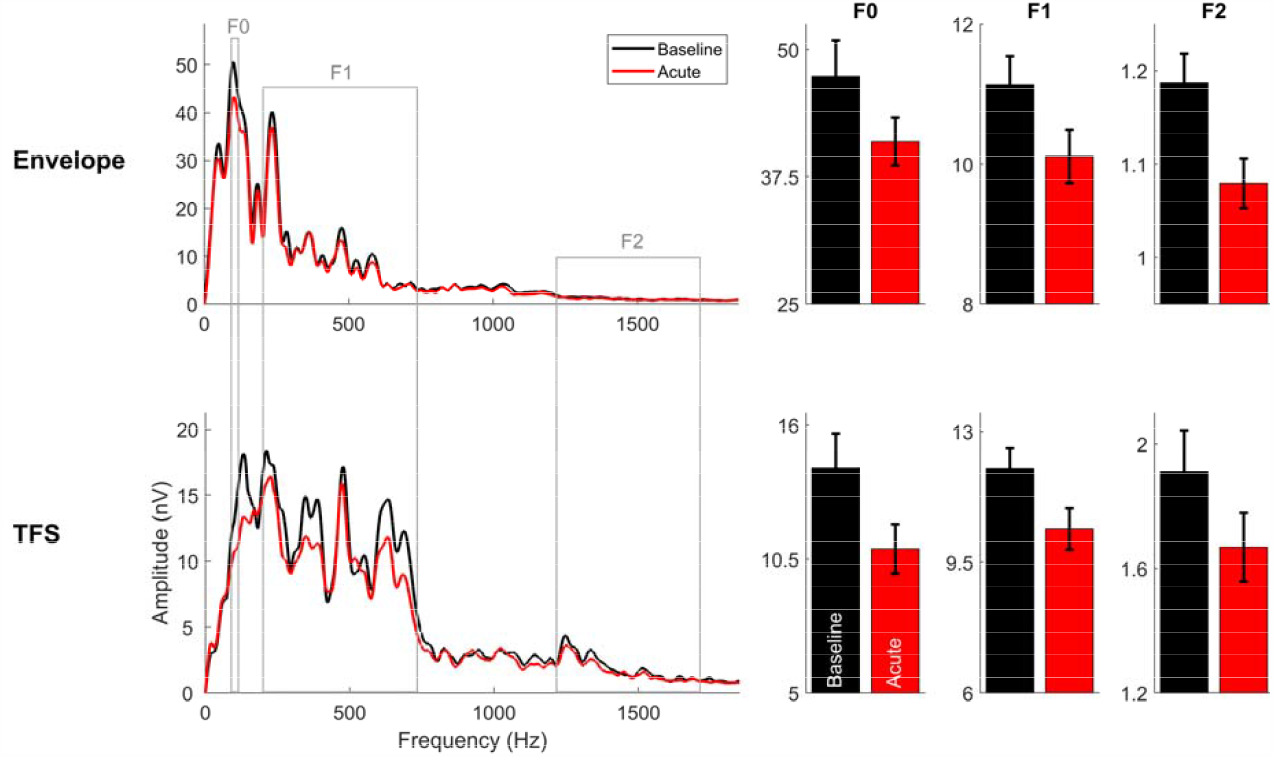
Spectral encoding. FFRs for student-athletes at baseline (black) and concussion (red) demonstrate smaller encoding post-concussion (left FFTs), with the differences in F0 being most pronounced in the envelope response (top FFT) and the F1 and F2 differences being most evident in the TFS (bottom FFT). Bar graphs (right) show the mean +1 standard deviation of the F0, F1, and F2 over these responses.

Consistent with previously established effects (38), there was a significant effect of polarity (added larger than subtracted responses; F_(1, 38)_ = 64.323, p < .001, η_p_ ^2^ = 0.629; Fig. 1) and frequency (F0 larger than F1 and F2; F_(2, 76)_ = 1719.115, p < .001, η _p_^**2**^ = 0.978; Fig. 1) and a polarity by frequency interaction (F0 largest in added, F2 largest in subtracted; F_(1, 38)_ = 218.334, p < .001, η _p_^2^ = 0.852; Fig. 1). The time by polarity, time by frequency, and time by polarity by frequency interactions were not significant (all p’s >.2), indicating that concussion results in an acute system-wide and feature-wide decline in auditory processing, rather than targeting any specific aspects of auditory processing.

### 3.1 FFR F0: Pitch Encoding

Planned comparison t-tests between acute and baseline for the envelope F0 showed a significant decline post-concussion (t_38_ = 2.143, p = .039, d = .343, means and standard deviation for all frequency regions provided in Table 1, Fig. 1), while this was only trending for the TFS response (t_38_ = 2.0, p = .053, d = .320, Fig. 1). The average reduction in envelope F0 amplitude was 6.4 ± 1.6 nV, which translates to an average percent change of -13.5%. For the TFS, the average decline in F0 amplitude was 3.3 ± 9.85 nV. This corresponds to, on average, a percent change of -23.3%. Of the 39 cases, 74.4% showed a decrease in envelope F0, while 64.1% of student-athletes showed a decrease in TFS F0 amplitude (Fig. 2).

**Table 1.** Mean and standard deviation (in nV) amplitude of F0, F1, and F2 for the envelope (env.) and temporal fine structure (TFS) responses at baseline (left) and acute (right) testing sessions.

|  |  | Baseline |  | Acute |  |
| --- | --- | --- | --- | --- | --- |
|  |  | Mean | SD | Mean | SD |
| **F0** | **Env.** | 47.36 | 21.93 | 40.98 | 14.68 |
|  | **TFS** | 14.23 | 8.89 | 10.92 | 6.32 |
| **F1** | **Env.** | 11.13 | 2.56 | 10.11 | 2.36 |
|  | **TFS** | 12.01 | 3.41 | 10.4 | 3.47 |
| **F2** | **Env.** | 1.19 | 0.2 | 1.08 | 0.17 |
|  | **TFS** | 1.91 | 0.84 | 1.67 | 0.69 |

**Figure 2.**
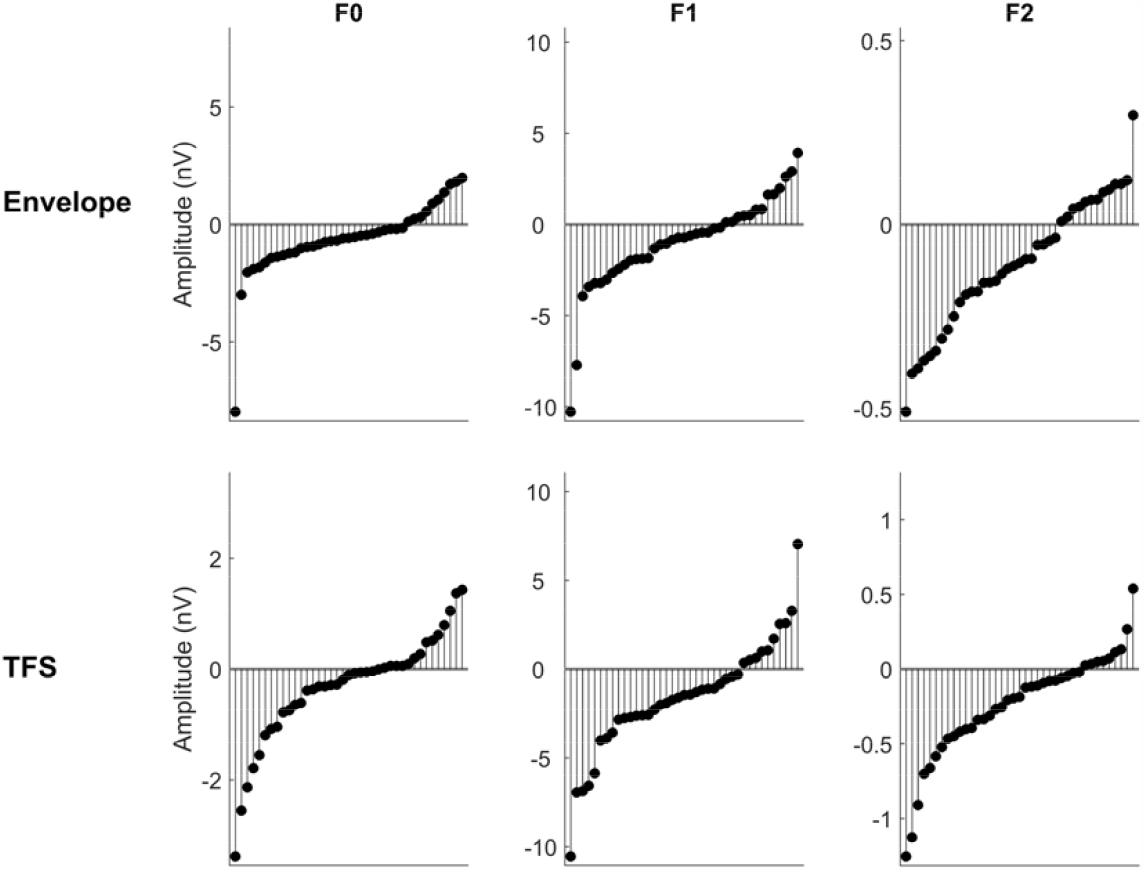
Difference plots for all individuals across all measures. Each plot shows the amplitude difference (in nV) between baseline and acute FFRs. A negative line indicates a decrease in encoding after a concussion. In both the envelope (top) and TFS (bottom) response, the majority of studentathletes had a decrease in encoding of the F0 (left), F1 (middle), and F2 (right).

### 3.2 FFR F1 and F2: Phonetic Encoding

#### 3.2a F1

Both envelope (t_38_ = 2.709, p = .01, d = .434, Fig. 1) and TFS (t_38_ = 3.296, p = .002, d = .528, Fig. 1) encoding of F1 declined following a concussion. The average decline in envelope F1 amplitude was 1.02 ± 2.63 nV, translating to an average percent change of -9.2%. For F1 amplitude of the TFS, there was an average decrease of 1.61 ± 3.16 nV, and a percent change of -13.4%. Of the 39 cases, 66.7% declined in envelope F1 encoding and 71.8% of cases decreased F1 encoding in the TFS following a concussion (Fig. 2).

#### 3.2b F2

Envelope (t_38_ = 3.566, p < .001, d = 0.571, Fig. 1) and TFS (t_38_ = 3.821, p < .001, d = 0.612, Fig. 1) encoding of F2 declined from baseline to acute testing. For envelope F2, the average decline of 0.11 ± 0.18 nV corresponds with a percent decrease of -9.2% and, for the F2 in the TFS, the average decline was 0.242 ± 0.358 nV, which corresponds to an average percent change of - 12.6%. For envelope F2 amplitude, 66.7% of student-athletes declined following a concussion and 76.9% of them showed a decline in TFS F2 amplitude (Fig. 2).

### Players with two concussions

At their second concussion, these 6 student-athletes once again showed a significant decline in frequency encoding following their injury (F_(1,5)_ = 8.473, p = .033, η _p_ ^2^ = .629, all means and standard deviations provided in Table 2; Fig. 3).

**Table 2.**
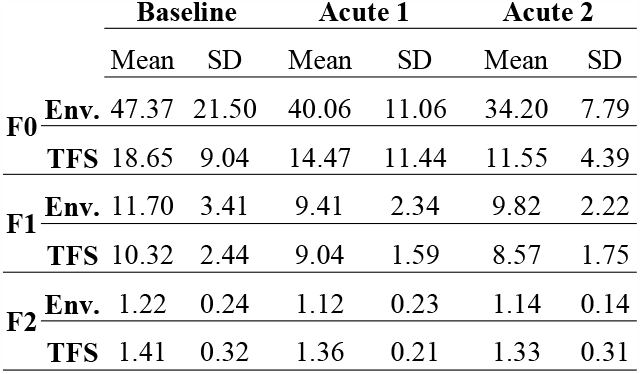
Means and standard deviations (in nV) at baseline, 1^st^ concussion, and 2^nd^ concussion for 6 student-athletes who sustained 2 concussions.

|  |  | Baseline |  | Acute 1 |  | Acute 2 |  |
| --- | --- | --- | --- | --- | --- | --- | --- |
|  |  | Mean | SD | Mean | SD | Mean | SD |
| **F0** | **Env.** | 47.37 | 21.50 | 40.06 | 11.06 | 34.20 | 7.79 |
|  | **TFS** | 18.65 | 9.04 | 14.47 | 11.44 | 11.55 | 4.39 |
| **F1** | **Env.** | 11.70 | 3.41 | 9.41 | 2.34 | 9.82 | 2.22 |
|  | **TFS** | 10.32 | 2.44 | 9.04 | 1.59 | 8.57 | 1.75 |
| **F2** | **Env.** | 1.22 | 0.24 | 1.12 | 0.23 | 1.14 | 0.14 |
|  | **TFS** | 1.41 | 0.32 | 1.36 | 0.21 | 1.33 | 0.31 |

**Figure 3.**
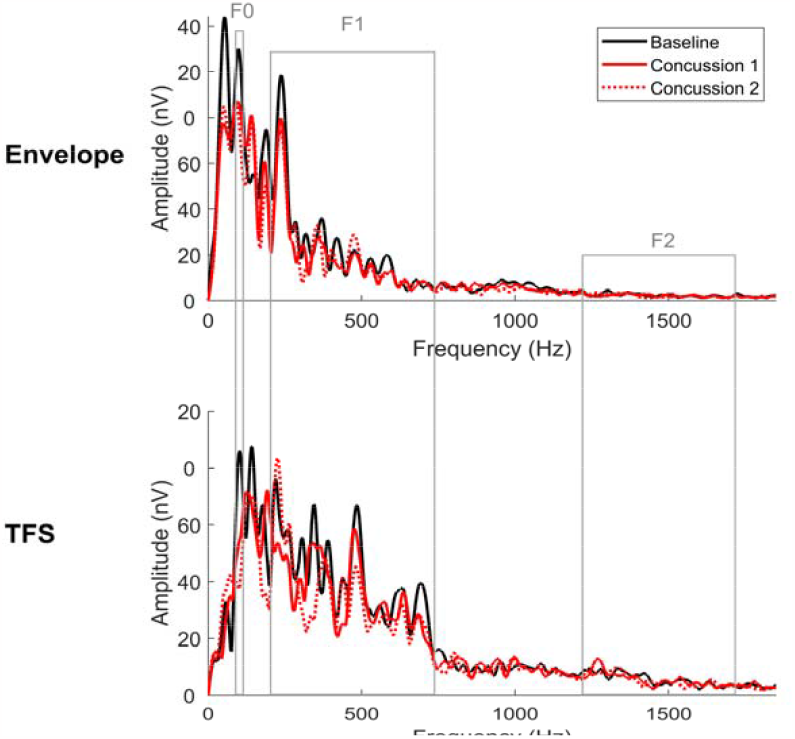
Test-retest FFTs. Averaged baseline FFTs (black) are plotted in comparison to the 1st (red solid) and 2nd (red dashed) concussions for the 6 individuals who had two injuries over the course of the study. FFTs are plotted for both the envelope (top) and temporal fine structure (TFS, bottom) responses. F0, F1, and F2 regions are indicated by a gray box. Note the similarity in FFR decline over F0, F1, and F2 in both the envelope and TFS following each concussion.

Again, as anticipated, there were significant effects of polarity (F_(1,5)_ = 24.910, p = .004,η _p^2^_ = .833) and frequency (F_(2,10)_ = 357.692,p < .001, _η p ^2^_ = .986), as well as a polarity by frequency interaction (F_(2,10)_ = 27.021, p< .001, η _P ^2^_ = .844). The interactions between injury status and polarity, injury status and frequency, and the three-way interaction among the three variables were not significant (all p’s > .24). The majority of these student-athletes showed a consistent change in frequency-encoding between their two concussions, and for most of these student-athletes, they declined in F0, F1, and F2 encoding following both injuries (Table 3).

**Table 3.** Number of student-athletes that showed a decline following each of their concussions and the number that changed in a consistent direction from baseline after each concussion for the envelope (Env.) and temporal fine structure (TFS) responses.

|  |  | F0 | F1 | F2 |
| --- | --- | --- | --- | --- |
| Env. | Decline at conc. 1 | 4/6 | 5/6 | 4/6 |
|  | Decline at conc. 2 | 4/6 | 5/6 | 4/6 |
|  | Consistent Change | 5/6 | 6/6 | 5/6 |
| TFS | Decline at conc. 1 | 4/6 | 5/6 | 4/6 |
|  | Decline at conc. 2 | 5/6 | 5/6 | 3/6 |
|  | Consistent Change | 5/6 | 6/6 | 5/6 |

The magnitude of change was also highly consistent between the two injuries. The difference between their healthy baseline and each injury was relatively stable, although most of the correlations were not significant, likely due to the very small sample size (Table 4; Fig 4).

**Figure 4.**
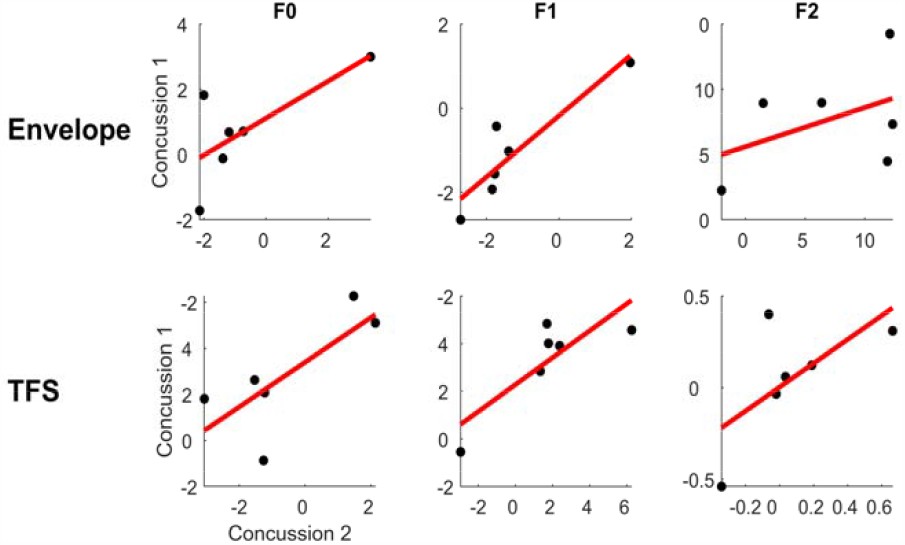
Scatterplots of envelope (top) and TFS (bottom) changes in F0 (left), F1 (middle), and HF (right) amplitude from baseline to each concussion for individuals with 2 concussions. The direction and magnitude of change from baseline for each FFR measure is highly consistent between the two concussions.

**Table 4.** **Correlation** and significance values comparing the change from baseline for the two concussions for the envelope (env.) and temporal fine structure (TFS) responses.

|  | F0 |  | F1 |  | F2 |  |
| --- | --- | --- | --- | --- | --- | --- |
|  | r | p | r | p | r | p |
| **Env.** | .720 | .107 | .918 | .012 | .446 | .376 |
| **TFS** | .755 | .083 | .833 | .04 | .658 | .156 |

## 4. Discussion

### 4.1 Central auditory system shows pervasive pathophysiology post-concussion

During the acute phase of concussion, the central auditory system shows a pervasive reduction in frequency encoding. For both the envelope and temporal fine structure responses, F0, F1, and F2 encoding are reduced following a head injury. The declines in pitch processing are consistent with findings in adolescents with post-concussion syndrome (32) and the long-term legacy of concussion (31) and subconcussion (34) in collegiate student-athletes. Considering both the previously reported effects together with the current findings of declines in both pitch and phonetic encoding identified in this study, we suggest that the auditory system demonstrates pervasive pathophysiology acutely following a sports-related concussion that partially recovers over time.

This auditory pathophysiology is objectively measurable using a non-invasive, simple test of auditory processing, namely the FFR. Given that the auditory system is susceptible to a concussion and that the FFR can provide an objective measure of concussion, consideration of adding the FFR to the multimodal assessment of sports-related concussion is warranted. Given the variability in type, location, and severity of contact that can induce a concussion, it is possible that adding a measure of auditory processing to the assessment may flag injuries that previously went unnoticed. By providing an objective window into auditory processing that has, to date, been overlooked, inclusion of the FFR could provide a more comprehensive snapshot of neural function in an athlete suspected of sustaining a sports-related concussion. The FFR may help the clinician weigh the subjective assessments reported by the patient together with the other neurosensory assessments that have become part of the standard protocol.

### 4.2 Understanding acute and long-term effects of concussion on auditory processing

In the context of previous findings in adolescents with post-concussion syndrome and recovered collegiate student-athletes (31, 32), these results support a distinction between acute and long-term effects of sports-related concussion on auditory processing. Together, these findings support a model where long-term effects are specific to pitch encoding, while, in the acute injury stage, auditory processing is pervasively affected, with both pitch and phonetic processing deficits observed. Given the differences observed at these two time points, we suggest that processing of pitch and phonetic information are both affected by a concussion but they recover along two different trajectories. The separate time courses of recovery for pitch and phonetic encoding could result from three possible mechanisms.

First, the observed differences in shortand long-term pathophysiology may be due to injury severity, whereby pitch encoding is impaired only in more severe injuries while phonetic encoding is always affected acutely following concussion. Encoding of phonetic information requires greater neural precision than pitch encoding because phonetic information is carried in higher frequencies. This precision depends on the balance of excitatory and inhibitory neurotransmitters released at synapses and integrated with neurons (14, 39, 40). Because a metabolic cascade of dysfunction upsets this communication between neurons (3, 5) and can lead to axonal dysfunction (4, 24, 25), it can presumably upset the precise neural timing required to process high-frequency, phonetic information. On the other hand, signals that encode pitch can be summed over a much broader temporal window (41, 42). A more deleterious injury that results in the death of axons through axonal shearing or the inability of a large number of neurons to regulate cell signaling may be necessary for disruptions in pitch processing to be observed. Measuring FFRs in individuals across a range of TBIs can test this hypothesis.

A second possibility is that the protracted pitch recovery is the result of auditory-system plasticity. That is, following a concussion, intact neurons alter signal encoding to account for the processing lost as a result of the injury. Pitch information is redundant in a speech signal and is thus encoded in multiple ways within the auditory system (43, 44). Given the higher proportion of auditory pathways devoted to pitch relative to phonetic encoding, it is likely that more pitch-encoding pathways would remain intact following a concussion. Thus, some of these pitch pathways may shift to phonetic encoding, to salvage encoding of the higher-frequency cues (45). To test this hypothesis, stimuli with pitches that are higher or lower in frequency to the current one could be used to determine if the same pitch and phonetic encoding changes are evident across frequencies.

Lastly, there is evidence to suggest that the higher-frequency components of the FFR represent signaling localized to subcortical circuits (i.e., the inferior colliculus) but that the lower-frequency pitch used in the current study may reflect pitch encoding across multiple cortical and subcortical auditory processing centers (29, 46-48). Thus, it is possible that acutely following the head injury there is neuroinflammation and axonal shearing that upsets the precise signaling between neighboring neurons required for high-frequency encoding but that recovery from these localized injuries occurs with time. In contrast, the response recorded to the pitch of the stimulus may show lasting impairments because it reflects across-brain health. That is, the long-term effects of concussion on pitch processing may reflect permanent, minor damage to axons at multiple centers along the auditory pathway, which sums in the scalp-recorded FFR into an observable deficit. This would suggest that pitch processing, as measured by the FFR may be a highly sensitive method of detecting subtle processing declines. This hypothesis is consistent with similar findings of a pitch-processing decline in a small group of concussed student-athletes that has been recently observed (33). Using a speech sound with a higher-frequency pitch to record FFRs in concussed student-athletes could test this hypothesis.

### 4.3 Auditory processing as an index of long-term pathophysiology

There is a real need to understand the link between repeat traumatic head injuries (rTBIs) and chronic traumatic encephalopathy. Currently, there is only a presumed link between rTBIs and CTE that is based on the high number of former collision athletes (e.g., football players or boxers) that show the tauopathy pathognomic of CTE (23, 49, 50). Given that the auditory system shows acute impairments that recover, but perhaps not fully, following a concussion, we think that long-term auditory pathophysiology, as measured by the FFR, may be a herald of subsequent CTE. In support of this, we see a similar impairment in pitch processing in collegiate student-athletes with long term participation in a contact or collision sport, even in individuals who have never experienced a concussion(34). A longitudinal study that continues to track collision athletes beyond retirement is necessary to confirm that the FFR can provide an in-vivo index of CTE.

### 4.4 Future directions

This study is novel because it is one of the first to examine acute changes in auditory physiology 24-72 hours post-concussion. Using comparison to an individual’s baseline, acute impairments are observed in pitch and phonetic processing. Although we do not have a post-recovery measure to determine if the brains of these student-athletes fully recovered to their preseason baseline, results from this study are nonetheless intriguing and motivate future studies and assessment of acute changes in auditory processing in more concussion cases. Importantly, subsequent studies need to consider concussion cases arising through other sports as well as determine if this acute auditory pathophysiology manifests in concussed females. We aim to continue testing any student-athletes who suffer a sports-related concussion to determine the generalizability of our findings across sports and sexes.

While these acute findings were observed following a *sports-related* concussion, we believe that they will be important in the assessment, diagnosis, and management of TBIs sustained through other head impacts, including blast exposure or accidents, such as car accidents or falls. This is an especially important consideration for the military as treatment of TBI in the military shares a similar goal of screening and treating injured individuals to return them to the contact-risk activity. A more comprehensive assessment can help ensure that the brain has fully healed. Future work could use the FFR to identify whether similar acute changes in auditory processing are observed following these brain injuries.

## Data Availability

All data produced in the present study are available upon reasonable request to the corresponding author at.

## Author Contributions

**JK:** Conceptualization, Methodology, Data Curation, Writing-Original Draft, Supervision, Project Administration; **DC:** Conceptualization, Methodology, Writing-Review & Editing; **JC:** Data Curation, Writing-Review & Editing; **SB:** Data Curation, Writing-Review & Editing; **TN:** Conceptualization, Methodology, Data Curation, Writing-Review & Editing; **MN:** Conceptualization, Methodology, Writing-Review & Editing; **NK:** Conceptualization, Methodology, Writing-Review & Editing, Supervision, Project Administration, Funding Acquisition

## 4.5 Conclusions

In conclusion, we find that the auditory system shows objective evidence of acute disruption following a sports-related concussion. These effects are pervasive; concussion impacts the fast encoding of the speech phonetics and the slower encoding of the voice pitch. The pattern of dysfunction was consistent across the majority of cases and motivates future studies that extend this work to other sports, other injuries (e.g., blast exposure), and to female athletes. It also suggests including the auditory system in the multimodal assessment of sports-related concussion. As an objective measure, the FFR can help provide a more complete picture of brain health in the acute and long-term assessment of athletes, especially those that participate in collision sports.

The authors thank members of the Auditory Neuroscience Laboratory, past and present, for their help with data collection. This research is funded by the National Institutes of Health under Grant: NIH R01-NS102500 and the Knowles Hearing Center, Northwestern University.

